# Somatic mosaic chromosomal alterations and death of cardiovascular disease causes among cancer survivors: an analysis of the UK Biobank

**DOI:** 10.1101/2022.08.20.22279019

**Authors:** Maxine Sun, Marie-Christyne Cyr, Johanna Sandoval, Louis-Philippe Lemieux Perreault, Lambert Busque, Jean-Claude Tardif, Marie-Pierre Dubé

## Abstract

Cancer survivors are at an increased risk of cardiovascular disease (CVD) compared to the general population. Here, we evaluated the impact of somatic mosaic chromosomal alterations (mCAs) on death of CVD causes, coronary artery disease (CAD) causes, from cancer, and of any cause in patients with a cancer diagnosis within the UK Biobank (n=48 919). mCAs were derived from DNA genotyping array intensity data and long-range chromosomal phase inference from participants. Overall, 10 070 individuals (20.6%) carried ≥1 mCA clone. In adjusted analyses, mCA was associated with an increased risk of death of CAD causes (hazard ratio [HR]: 1.37, 95% confidence interval [CI]: 1.09-1.71, *P*=0.006), from cancer (HR: 1.06, 95% CI: 1.00-1.11, *P*=0.041), and death of any cause (HR: 1.07, 95% CI: 1.02-1.12, *P*=0.005). Among cancer survivors, carriers of any mCA are at an increased risk of death of CAD causes and of any cause as compared to non-carriers.

## INTRODUCTION

The number of survivors of cancer is growing worldwide due to the ageing populations and improvements in early cancer detection and treatment modalities.^1^ It is estimated that over 26 million people in the United States alone will be living with a history of cancer by the year 2040.^2,3^ Among cancer survivors, a pressing clinical problem is their increased predisposition to cardiovascular disease (CVD)^4,5^ and treatment-related cardiac dysfunction^6,7^. Currently, there are no guideline recommendations with respect to CVD screening for patients with cancer, possibly stemming from the lack of CVD-related markers that can better risk-stratify cancer patients beyond existing cardiovascular risk factors for the general population.^8^ The identification and development of biomarkers that can eventually be used towards a risk assessment tool for the purpose of discriminating patients diagnosed with cancer who are at a higher risk of CVD may be useful.^8^

Clonal Hematopoiesis (CH) refers to a population of cells derived from a mutated multipotent stem/progenitor cell occurring in the context of aging.^9^ CH can be caused by somatic mutation in driver genes^10-14^ called clonal hematopoiesis of indeterminate potential (CHIP)^15^, or by somatic mosaic chromosomal alterations (mCA).^16-20^ Previously, CHIP has been associated with a greater burden of atherosclerotic vessel disease^21^, a higher risk of myocardial infarction^22,23^, inflammatory response^24^, and death of any cause^11,12^. The presence of CHIP has also been associated with treatment-related adverse outcomes in cancer survivors.^25,26^

Somatic mCAs correspond to large chromosomal gains, loss, or copy neutral losses of heterozygosity which can affect autosomes or sexual chromosomes.^27^ The most prevalent mCA is the loss of the Y chromosome in aging men.^28^ Mosaic loss of the Y chromosome has been associated with all-cause mortality^29,30^, Alzheimer’s disease^31^, autoimmune disease^32^, diabetes^29^, and cardiovascular events^33^.

Given the links between CH and CVD in the normal aging population, and the paucity of data in cancer survivors, we sought to evaluate the impact of mCAs on the risk of death from CVD causes, from cancer, and of any cause among cancer survivors at risk of CVD by relying on DNA genotyping array intensity data and long-range chromosomal phase information inferred from participants of the UK Biobank. Our hypothesis was that carriers of any mCA would be at higher risk of death from CVD causes compared to their mCA non-carriers.

## Results

### Baseline characteristics and mCA prevalence

Overall, 48 919 participants were diagnosed with bladder, larynx, corpus uteri, prostate, rectal, breast, kidney, non-Hodgkin lymphoma, melanoma, or lung cancer within the UK Biobank (**Table 1**). The mean age at study entry was 60 years old (median 62, interquartile range [IQR]: 56-65). Forty-six percent (45.8%) were men and 39.6% previously smoked.

**Table 1.**
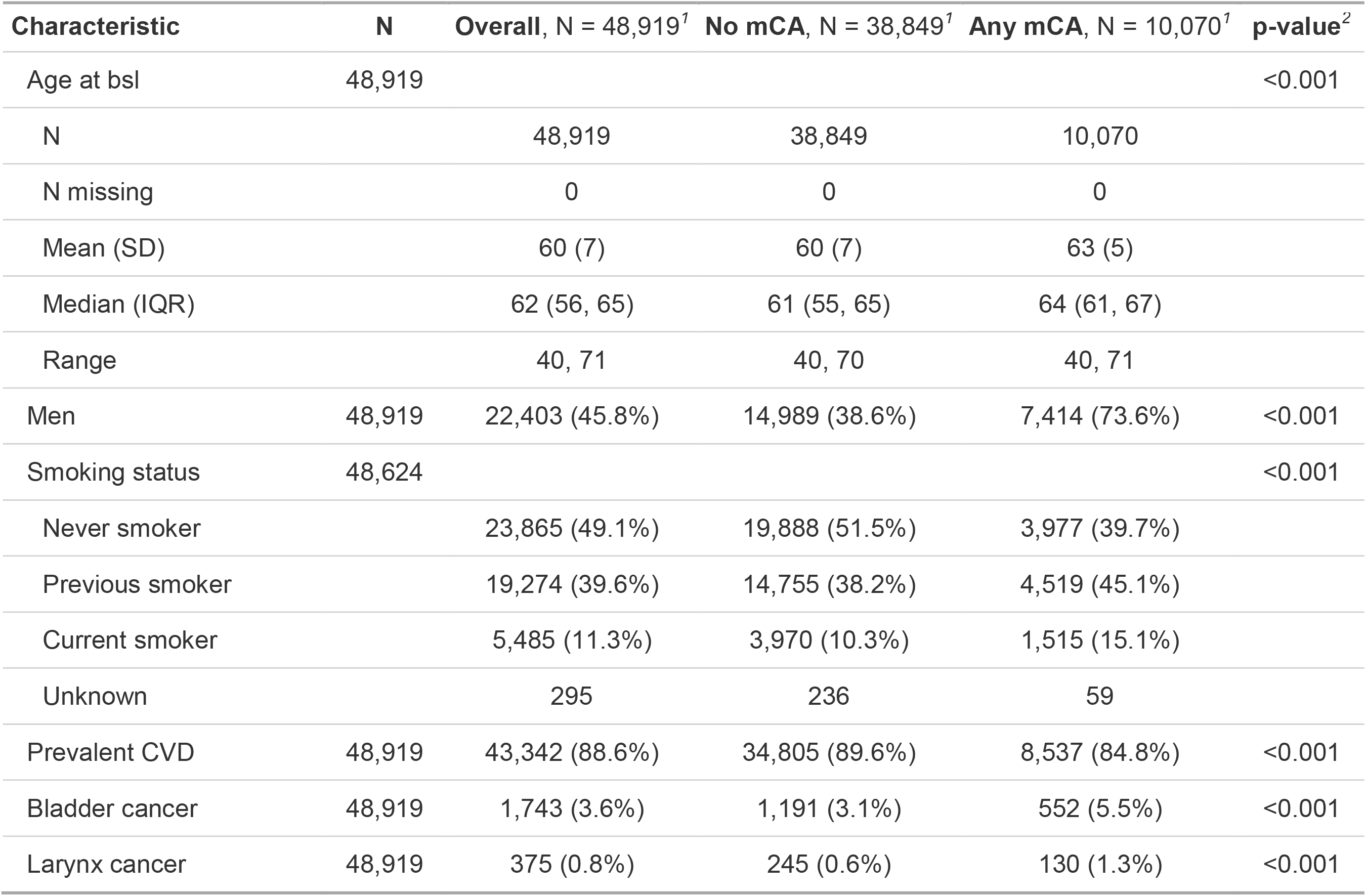
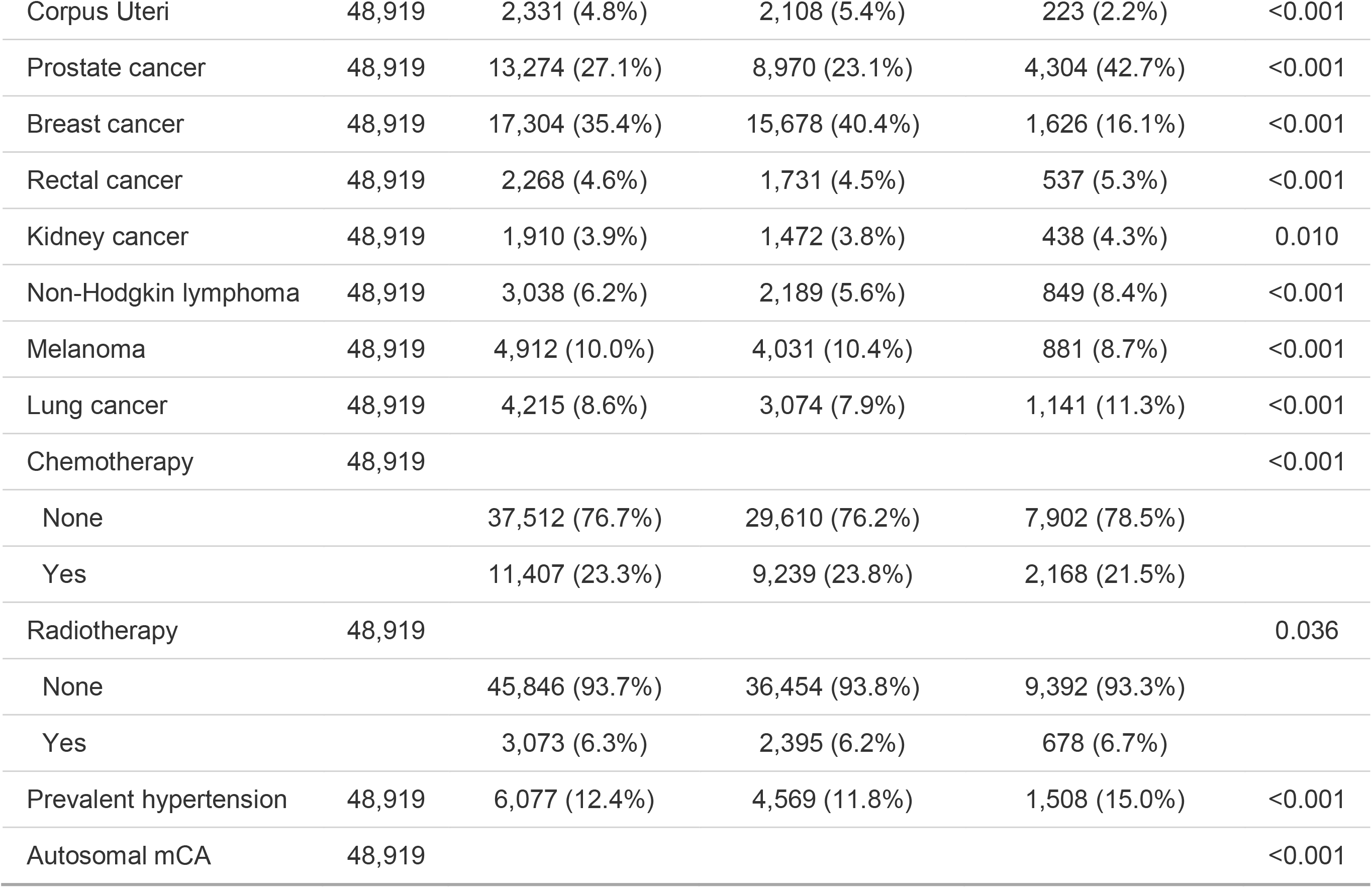
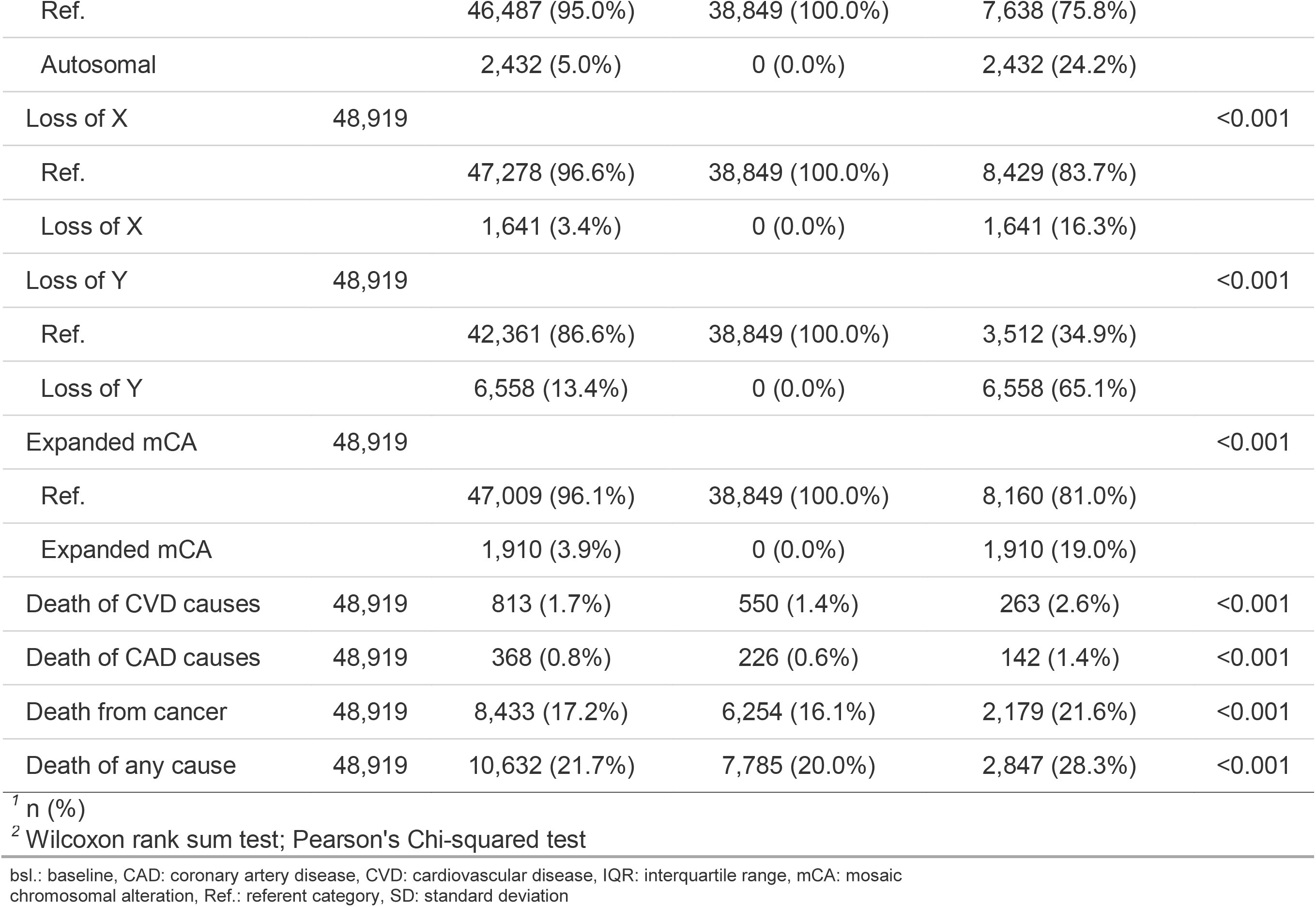
Summary descriptives of patients with a cancer diagnosis according to mosaic chromosomal alterations status. mCA: mosaic chromosomal alteration; IQR: interquartile range; SD: standard deviation; CVD: cardiovascular disease; CAD: coronary artery disease

| Characteristic | N | Overall, N = 48,919<sup>1</sup> | No mCA, N = 38,849<sup>1</sup> | Any mCA, N = 10,070<sup>1</sup> | p-value<sup>2</sup> |
| --- | --- | --- | --- | --- | --- |
| Age at bsl | 48,919 |  |  |  | <0.001 |
| N |  | 48,919 | 38,849 | 10,070 |  |
| N missing |  | 0 | 0 | 0 |  |
| Mean (SD) |  | 60 (7) | 60 (7) | 63 (5) |  |
| Median (IQR) |  | 62 (56, 65) | 61 (55, 65) | 64 (61, 67) |  |
| Range |  | 40, 71 | 40, 70 | 40, 71 |  |
| Men | 48,919 | 22,403 (45.8%) | 14,989 (38.6%) | 7,414 (73.6%) | <0.001 |
| Smoking status | 48,624 |  |  |  | <0.001 |
| Never smoker |  | 23,865 (49.1%) | 19,888 (51.5%) | 3,977 (39.7%) |  |
| Previous smoker |  | 19,274 (39.6%) | 14,755 (38.2%) | 4,519 (45.1%) |  |
| Current smoker |  | 5,485 (11.3%) | 3,970 (10.3%) | 1,515 (15.1%) |  |
| Unknown |  | 295 | 236 | 59 |  |
| Prevalent CVD | 48,919 | 43,342 (88.6%) | 34,805 (89.6%) | 8,537 (84.8%) | <0.001 |
| Bladder cancer | 48,919 | 1,743 (3.6%) | 1,191 (3.1%) | 552 (5.5%) | <0.001 |
| Larynx cancer | 48,919 | 375 (0.8%) | 245 (0.6%) | 130 (1.3%) | <0.001 |
| Corpus Uteri | 48,919 | 2,331 (4.8%) | 2,108 (5.4%) | 223 (2.2%) | <0.001 |
| Prostate cancer | 48,919 | 13,274 (27.1%) | 8,970 (23.1%) | 4,304 (42.7%) | <0.001 |
| Breast cancer | 48,919 | 17,304 (35.4%) | 15,678 (40.4%) | 1,626 (16.1%) | <0.001 |
| Rectal cancer | 48,919 | 2,268 (4.6%) | 1,731 (4.5%) | 537 (5.3%) | <0.001 |
| Kidney cancer | 48,919 | 1,910 (3.9%) | 1,472 (3.8%) | 438 (4.3%) | 0.010 |
| Non-Hodgkin lymphoma | 48,919 | 3,038 (6.2%) | 2,189 (5.6%) | 849 (8.4%) | <0.001 |
| Melanoma | 48,919 | 4,912 (10.0%) | 4,031 (10.4%) | 881 (8.7%) | <0.001 |
| Lung cancer | 48,919 | 4,215 (8.6%) | 3,074 (7.9%) | 1,141 (11.3%) | <0.001 |
| Chemotherapy | 48,919 |  |  |  | <0.001 |
| None |  | 37,512 (76.7%) | 29,610 (76.2%) | 7,902 (78.5%) |  |
| Yes |  | 11,407 (23.3%) | 9,239 (23.8%) | 2,168 (21.5%) |  |
| Radiotherapy | 48,919 |  |  |  | 0.036 |
| None |  | 45,846 (93.7%) | 36,454 (93.8%) | 9,392 (93.3%) |  |
| Yes |  | 3,073 (6.3%) | 2,395 (6.2%) | 678 (6.7%) |  |
| Prevalent hypertension | 48,919 | 6,077 (12.4%) | 4,569 (11.8%) | 1,508 (15.0%) | <0.001 |
| Autosomal mCA | 48,919 |  |  |  | <0.001 |
| Ref. |  | 46,487 (95.0%) | 38,849 (100.0%) | 7,638 (75.8%) |  |
| Autosomal |  | 2,432 (5.0%) | 0 (0.0%) | 2,432 (24.2%) |  |
| Loss of X | 48,919 |  |  |  | <0.001 |
| Ref. |  | 47,278 (96.6%) | 38,849 (100.0%) | 8,429 (83.7%) |  |
| Loss of X |  | 1,641 (3.4%) | 0 (0.0%) | 1,641 (16.3%) |  |
| Loss of Y | 48,919 |  |  |  | <0.001 |
| Ref. |  | 42,361 (86.6%) | 38,849 (100.0%) | 3,512 (34.9%) |  |
| Loss of Y |  | 6,558 (13.4%) | 0 (0.0%) | 6,558 (65.1%) |  |
| Expanded mCA | 48,919 |  |  |  | <0.001 |
| Ref. |  | 47,009 (96.1%) | 38,849 (100.0%) | 8,160 (81.0%) |  |
| Expanded mCA |  | 1,910 (3.9%) | 0 (0.0%) | 1,910 (19.0%) |  |
| Death of CVD causes | 48,919 | 813 (1.7%) | 550 (1.4%) | 263 (2.6%) | <0.001 |
| Death of CAD causes | 48,919 | 368 (0.8%) | 226 (0.6%) | 142 (1.4%) | <0.001 |
| Death from cancer | 48,919 | 8,433 (17.2%) | 6,254 (16.1%) | 2,179 (21.6%) | <0.001 |
| Death of any cause | 48,919 | 10,632 (21.7%) | 7,785 (20.0%) | 2,847 (28.3%) | <0.001 |

<sup>1</sup> n (%)
<sup>2</sup> Wilcoxon rank sum test; Pearson's Chi-squared test
bsl.: baseline, CAD: coronary artery disease, CVD: cardiovascular disease, IQR: interquartile range, mCA: mosaic chromosomal alteration, Ref.: referent category, SD: standard deviation

The presence of at least one mCA clone was observed in 20.6% (10 070/48 919) of patients **(Suppl. Materials, sFigure 1A)**. Of those, 2 432 were autosomal carriers and 1 910 had mCA in ≥10% of peripheral leukocytes, defined as an expanded mCA clone. Whereas most carriers had only one mCA, 812 individuals carried ≥2 non-overlapping mCAs. In general, those with mCA were older (median 64 vs. 61 years), where the prevalence of mCA increased with increasing age categories: 4.8% among those aged <49 years old, 12.1% for those aged 50-59 years old, 26.4% for those aged 60-69 years old, and 30.4% for those aged ≥70 years old (**Suppl. Materials, sFigure 1B**). Expectedly, 73.6% of patients with mCA were men, likely because the majority of mCA carriers was due to loss of the Y chromosome (n=6 558). In comparison, mCA due to loss of the X chromosome was observed in 1 641 patients (16.3%). Amongst patients older than 65 years old, the prevalence of mCA according to cancer types was the lowest for breast cancer (14%) and the highest for larynx cancer (43%, **Suppl. Materials, sFigure 1C**).

### mCA and the risk of death of CVD causes, coronary artery disease (CAD) causes, from cancer, and any cause

In adjusted Cox regression analyses, mCA was associated with an increased risk of death of coronary artery disease (CAD) causes (hazard ratio [HR]: 1.37, 95% confidence interval [CI]: 1.09-1.71, *P*=0.006), death from cancer (HR: 1.06, 95% CI: 1.00-1.11, *P*=0.041), and death of any cause (HR: 1.07, 95% CI: 1.02-1.12, *P*=0.005, **Figure 1**). These effects were maintained after the additional adjustment for hypertension and body-mass index in sensitivity analyses (**Suppl. Materials, sTable 1**). Sub-analyses according to smoking status showed that mCAs were associated with significantly higher risks of death of CVD causes (smoking-by-mCA *P*_interaction_=0.044), and death of any cause (smoking-by-mCA *P*_interaction_=0.011), but not with death of CAD-causes (smoking-by-mCA *P*_interaction_=0.50) or with death from cancer (smoking-by-mCA *P*_*interaction*_=0.266, **Suppl. Materials, sTable 2**). In sub-analyses considering chemotherapy, chemotherapy-by-mCA interaction terms were not significant for any of the primary endpoints (all *P*_interaction_≥0.05, **Suppl. Materials, sTable 3**).

**Figure 1.**
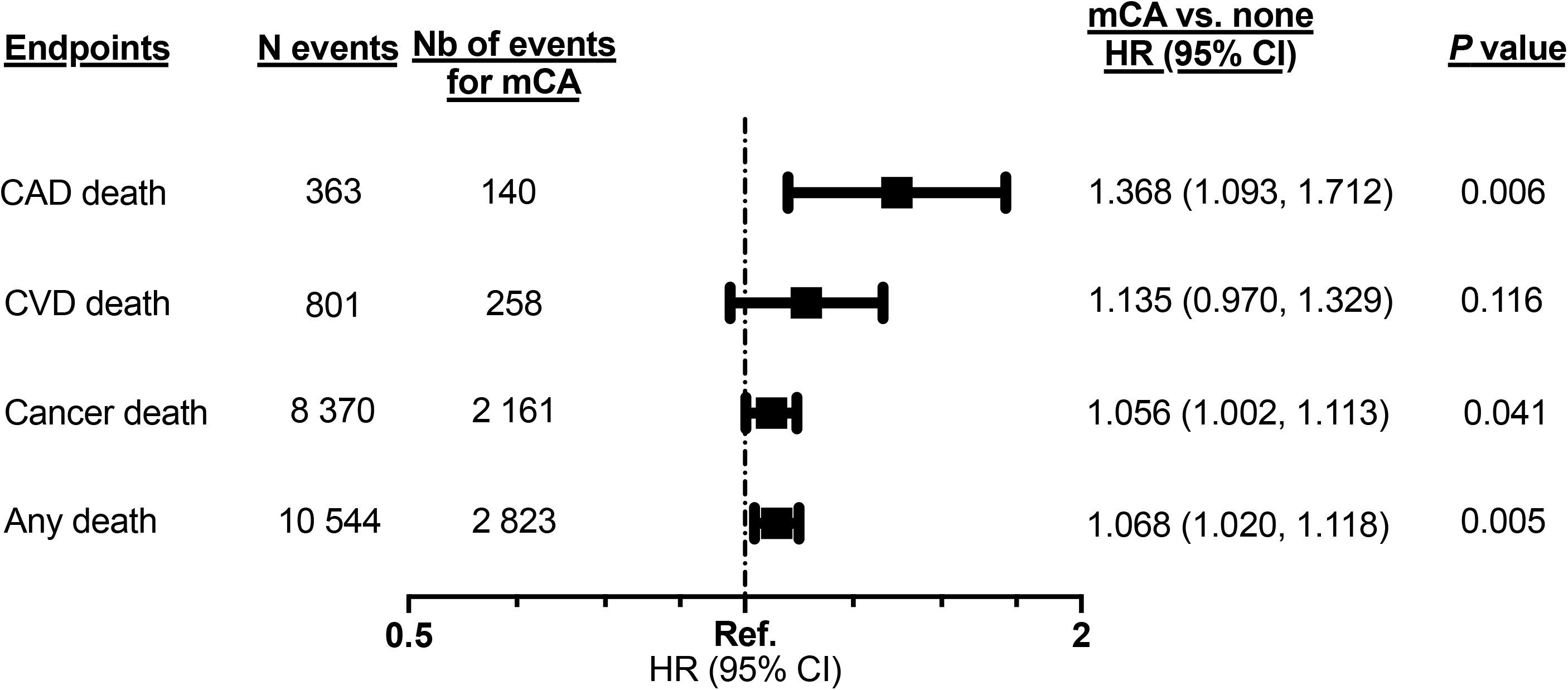
Multivariable Cox regression models evaluating the effect of mosaic chromosomal alterations on death of cardiovascular disease causes, of coronary artery disease causes, from cancer, and death of any cause. All models are adjusted for age at baseline, sex, smoking status, chemotherapy, radiotherapy, number of days between the date of recruitment and the date of cancer diagnosis, and principal components 1 to 10. CAD: coronary artery disease, CI: confidence interval, CVD: cardiovascular disease, HR: hazard ratio, mCA: mosaic chromosomal alterations, Ref.: referent category (1.0)

Different types of mCAs had different effects on the study endpoints. Notably, autosomal mCAs were associated with death of CVD causes (HR: 1.35, 95% CI: 1.04-1.75, *P*=0.023), with death from cancer (HR: 1.14, 95% CI: 1.04-1.24, *P*=0.004), as well as with death of any cause (HR: 1.14, 95% CI: 1.06, 1.24, *P*<0.001, **Suppl. Materials, sTable 4**). mCA due to loss of the X chromosome was associated with death of CAD causes (HR: 2.02, 95% CI: 1.16, 3.54, *P*=0.013, **Suppl. Materials, sTable 5**), and mCA due to loss of the Y chromosome was associated with death of any cause (HR: 1.08, 95% CI: 1.02-1.14, *P*=0.009, **Suppl. Materials, sTable 6**). Expanded mCAs taken individually had no statistically significant impact on the primary endpoints (**Suppl. Materials, sTable 7**). When considering the study population of 479 435 participants with, and without a history of any cancer, the interaction terms between cancer and mCA were significant for death from cancer (*P*_interaction_<0.001) and death of any cause (*P*_interaction_<0.001, **Suppl. Materials, sTable 8**). On the other hand, the interaction terms between cancer status and mCA for death of CVD causes and CAD causes were not statistically significant (**Suppl. Materials, sTable 8**).

### mCA and the risk of death of CVD causes, CAD causes, from cancer, and any cause according to cancer site

In multivariable analyses where the effect of mCA was assessed in subgroups according to cancer type, we found that carriers of mCAs diagnosed with kidney cancer had an increased risk of death of CVD causes (HR: 2.03, 95% CI: 1.11-3.72, *P*=0.022) and CAD causes (HR: 3.57, 95% CI: 1.44-8.84, *P*=0.006, **Suppl. Materials, sTables 9 & 10**) compared to their mCA non-carriers counterparts diagnosed with kidney cancer. Women diagnosed with breast cancer who carried a mCA also had a higher risk of death of CAD causes (HR: 2.46, 95% CI: 1.23-4.92, *P*=0.011). mCA had no significant impact on deaths from cancer in either cancer type (**Suppl. Materials, sTable 11**). Women carriers of mCA diagnosed with corpus uteri cancer had an increased risk of death of any cause (HR: 1.35, 95% CI: 1.01-1.80, *P*=0.042, **Suppl. Materials, sTable 12**).

### mCA and the risk of incident CV endpoints (exploratory analyses)

In the overall cohort of cancer survivors (n=48 919), carriers of mCA had a higher risk of incident ST-elevation myocardial infarction (STEMI) (HR: 1.19, 95% CI: 1.03-1.37, *P*=0.022) and peripheral vascular disease (HR: 1.17, 95% CI: 1.04-1.31, *P*=0.007, **Figure 2**) than mCA non-carriers. Amongst those aged 65 years and above, carriers of mCA showed a higher risk of incident STEMI (HR: 1.23, 95% CI: 1.02-1.49, *P*=0.034), non-STEMI (HR: 1.20, 95% CI: 1.03-1.40, *P*=0.020), stable angina (HR: 1.20, 95% CI: 1.00-1.44, *P*=0.047), intracerebral hemorrhage (HR: 1.22, 95% CI: 1.01-1.48, *P*=0.038), subarachnoid hemorrhage (HR: 1.23, 95% CI: 1.01-1.50, *P*=0.040), and peripheral vascular disease (HR: 1.20, 95% CI: 1.03-1.39, *P*=0.017, **Suppl. Materials, sTable 13**).

**Figure 2.**
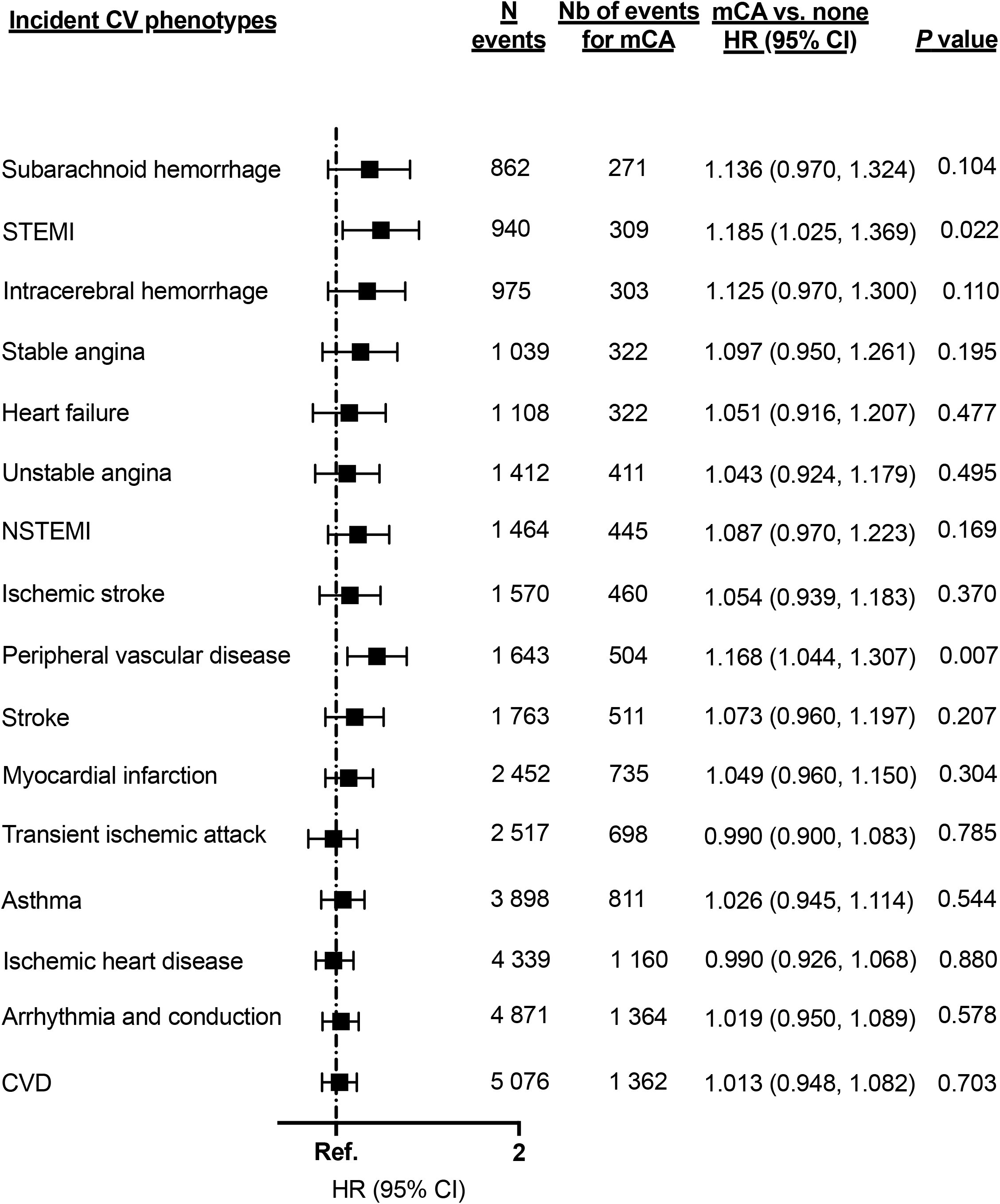
Multivariable Cox regression models evaluating the effect of mosaic chromosomal alterations on incident cardiovascular phenotypes. All models are adjusted for age at baseline, sex, smoking status, chemotherapy, radiotherapy, number of days between the date of recruitment and the date of cancer diagnosis, and principal components 1 to 10. CVD: cardiovascular disease, CI: confidence interval, HR: hazard ratio, mCA: mosaic chromosomal alteration, MI: myocardial infarction, NSTEMI: non-ST-elevation myocardial infarction, STEMI: ST-elevation myocardial infarction, TIA: transient ischemic attack.

## DISCUSSION

Here, we assessed the impact of somatic mCAs, a subtype of CH, on the risks of death of CVD causes, CAD causes, from cancer, and death of any cause focusing exclusively on individuals diagnosed with one of 10 cancer types known to have the highest reported rates of CVD deaths^34^. First, we found that carriers of mCAs had an increased risk of death of CAD causes, from cancer, and of any cause. Some of the associations were more impactful in mCA subtypes, notably autosomal mCAs were associated with the risk of death of CVD causes, mCA due to loss of the X chromosome was associated with the risk of death of CAD causes, and mCA due loss of the Y chromosome was associated with the risk of death of any causes.

Second, exploratory analyses suggest that carriers of mCAs were at higher risk of incident STEMI and peripheral vascular disease; and for cancer survivors aged >65 years, at a higher risk of incident NSTEMI, stable angina, intracerebral hemorrhage, and subarachnoid hemorrhage. In comparison, a previous mCA study focusing on MI and stroke did not find any significant associations when focusing on carriers of expanded mCAs, regardless of cancer status.^20^ The current study suggests that any mCA carrier status can be prognostic of several other CV phenotypes in cancer survivors. Previous research looking at mutation driven CH have correlated mutations in *DNMT3A* and *TET2* with chronic heart failure in patients with STEMI (n=485).^35^ In another study including approximately 50 000 samples with exome sequencing data, Zekavat et al.^26^ found that carriers of CHIP mutations had higher risks of peripheral artery disease. Third, when considering cancer survivors, it is possible that cancer-directed treatments may have perpetuated the influence of mCA on outcomes.^25^ However, our sub-analyses which included a chemotherapy-by-mCA interaction term failed to reach statistical significance, albeit a near 2-fold increased risk of death of CAD causes for mCA carriers. Previous work have supported the hypothesis that chemotherapy may lead to clonal expansion of mutations in apotosis or DNA repair gens (*TP53, PPM1D*)^25,36^, and that CHIP mutations caused by these genes could worsen the risk of atherosclerosis^26^. It is possible that chemotherapy usage may not be well-captured in the current database. Furthermore, without knowledge on tumor aggressiveness in the current database, it was not possible to exclude the presence of a selection bias where carriers of mCAs may be at a more advanced stage of the disease or have a more aggressive tumor histology, which may make them more likely to undergo additional lines of chemotherapy, which could have contributed to their risks of CVD. It will be critical to evaluate whether cancer-directed treatments have an additive or synergistic influence on mCAs. Further research will be needed to understand the impact of mCAs on CVD outcomes following modern standards of care, such as treatment with immune-checkpoint inhibitors. In that scenario, mCA carriers may benefit from an in-depth preventive cardiology approach prior to treatment where multi-disciplinary cardio-oncology care could mitigate such risks.

Fourth, we found that the effect of mCA on the risks of death of CVD and CAD causes, as well as of death of any cause was specific to previous smokers. This upholds the findings of a recent study which established strong causal associations between smoking and mCAs, and postulated that smoking may contribute to the selection of clones bearing somatic mutations.^37^ Indeed, we found that cancer survivors who are carriers of mCAs may face added risks if they were previous smokers. Additional work is required to better evaluate how smoking habits can impact mCAs and the downstream consequences of such mutations, especially in patients diagnosed with cancer.

Nevertheless, the generalizability of our findings may be limited as the influence of mCA was not consistent across all examined cancer types. This was not entirely unexpected given the heterogeneous biology of cancers included and distinct mechanisms of mCAs with respect to these cancers and treatments that patients received. Even so, the associations observed within this study may inform future studies focusing on the effect of mCA and the risks of death from CVD causes for specific cancer types.

Our study has additional limitations. First, our study did not consider CHIP mutations. While the association of CHIP mutations with CVD has been established, it has not been formally tested in patients diagnosed exclusively with non-hematologic cancer. There is reason to believe that carriers of CHIP mutations in addition to being diagnosed with cancer may face higher risks of CVD. In addition, the synergistic effect of both mCA and CHIP mutation carriers amongst cancer survivors will be of interest to explore in future studies. Second, It has been established that detectable hematopoietic loss of the X and Y chromosomes increase with age, with more age-related pathologies in older individuals than in younger individuals.^38,39^ Unexpectedly, the association between the mosaic loss of the Y chromosome and death of CVD causes was not significant in the current analysis that focused on cancer survivors. The non-significant association may have been related to mosaic loss of the Y chromosome definition used, where a larger percentage threshold of blood cells lacking chromosome Y may be more relevant. For example, a recent analysis found that regardless of their cancer status, men with mosaic loss of the Y chromosome in 40% or more of leukocytes displayed a 31% increased risk of dying from any disease related to the circulatory system based on survival data from the UK Biobank.^39^ In contrast, the relative risk per 1% increase of detectable loss of Y chromosome and the risk of death from CVD causes was more subdued (HR: 1.0054, *P*=0.001).^39^ We did also note that mosaic loss of the Y chromosome was rather impactful for cancer-specific mortality, which may be relevant in considering the significant association found between loss of the Y chromosome and death of any cause. Third, mCA status was evaluated only at study entry. Reasonably, clones carrying a mCA may undergo rapid changes following various environmental exposures (e.g. smoking, chemotherapy). Under this premise, serial measurements of mCA may better help inform the dose-response effect of such factors and mCA. Finally, while adjustment was made for chemotherapy and radiotherapy treatment, some individuals may have undergone such treatments in the outpatient setting, which would not have been properly captured using inpatient procedural codes.

In conclusion, our study showed that mCAs are associated with higher risks of death of CAD causes among cancer survivors. mCAs were also associated with higher risks of death from cancer and death of any cause. Notably, autosomal mCAs appeared to be driving most of the observed associations. Future studies may focus on specific cancer types and their treatments to better ascertain the effect of mCA on the risk of CVD and evaluate if they constitute a useful biomarker in the management of cancer survivors.

## METHODS

### Study samples

The UK Biobank is a large population-based cohort that includes over 500,000 participants aged between 40-70 years old, recruited from 2006 to 2010.^40^ Baseline interviews regarding their medical history and environmental exposures were conducted in the UK across 22 assessment centers where blood samples for genotyping were also obtained and blood analysis performed. Additional health outcome data, including diagnoses of cancer and CVD, have been linked via UK national registries and hospital records managed by the NHS, and genome-wide genotyping of blood-derived DNA was performed by the UK Biobank using 2 genotyping arrays sharing 95% of marker content: Applied Biosystems UK BiLEVE Axiom Array and Applied Biosystems UK Biobank Axiom Array, both by Affymetrix.^41^

### Determination of mosaic chromosomal alterations

Previously, Loh et al.^20^ identified mosaic chromosomal alteration in 487 409 genotyped individuals. As previously described^19,20^, mCAs were determined from genotype intensities log_2_R ratio (LRR) and B-allele frequency (BAF) values, which were used to estimate the total and relative allelic intensities, respectively. Re-phasing was conducted using Eagle2^42^ and mCA calling was performed by leveraging long-range phase information searching for allelic imbalances between maternal and paternal allelic fractions across contiguous genomic segments. For the purpose of our study, mCA calls were obtained from dataset Return 3094 from the UK Biobank application 19808.^20,42^ From the genetic data of the UK Biobank, 479 435 individuals who passed the sample quality control criteria, including genotypic-phenotypic sex concordance, and without first or second-degree relatives in the dataset were considered (**Figure 3**). Of those, we identified 48 919 participants with a diagnosis of cancer before or after the baseline assessment visit based on the cancer register, using ICD-9 and ICD-10 diagnostic codes for bladder, larynx, prostate, corpus uteri, rectum, breast, kidney, non-Hodgkin lymphoma, melanoma of the skin, or lung cancer (**Suppl. Materials, sTable 14)**. These specific cancer types were selected based on a previous publication which identified the top 10 cancer sites with the largest percentage of deaths attributed to CVD.^34^

**Figure 3.**
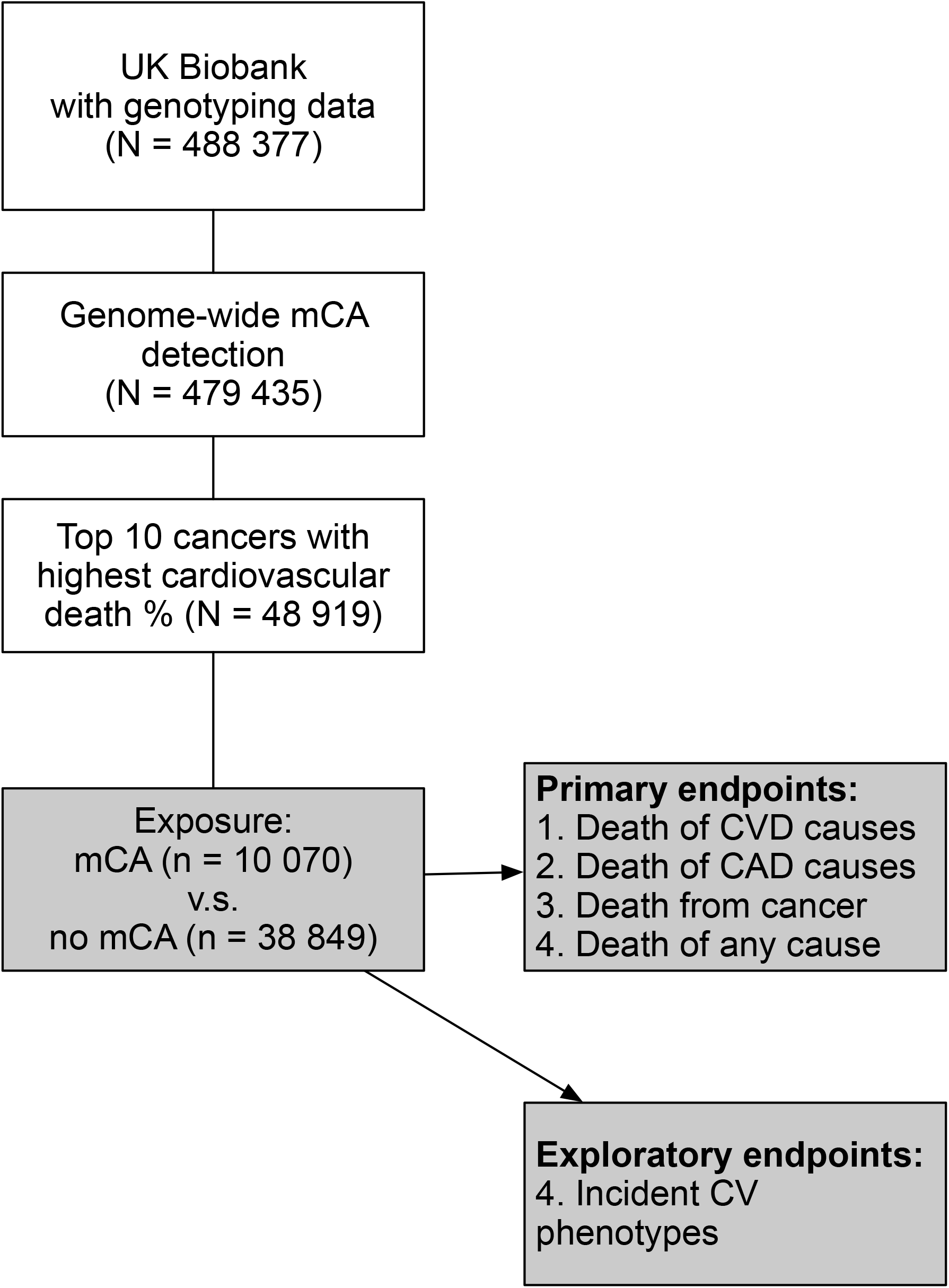
Visual representation of the study flow.

### Exposure and clinical outcomes

The exposure of interest was the presence of a mCA of any type, which was classified as ≥1 or none. mCAs were also categorized as autosomal mCAs, loss of the X chromosome, loss of the Y chromosome, and expanded mCAs (defined as mCAs present in at least 10% of peripheral leukocyte DNA indicative of clonal expansion^43^). The study primary endpoints consisted of death of CVD causes, death of CAD causes, death from cancer, and death of any cause based on ICD-10 codes for primary cause of death from the death register records (**Suppl. Materials, sTable 15**). For each endpoint, the time to death was calculated from the date of the assessment visit at the time of recruitment into the UK Biobank (baseline) if patients’ cancer diagnosis occurred *<u>before</u>* baseline, and alternatively, from the date of cancer diagnosis if the cancer diagnosis occurred *<u>after</u>* baseline. The number of days between a patient’s cancer diagnosis date and baseline was recorded, and it was set as 0 if the cancer diagnosis occurred after baseline. For individuals who were not deceased, the end of follow-up was the last date of death registered based on participant’s country of enrolment (i.e., 2021-03-30 for England, 2021-03-23 for Scotland and 2021-03-16 for Wales).

Other exploratory endpoints included various incident CV phenotypes^44^ recorded after patients’ cancer diagnosis date (**Suppl. Materials, sTable 15**). For these endpoints, the time to the event (i.e., incident CV event of interest or death of CVD causes) was calculated from the date of cancer diagnosis. For those who did not experience any incident CV events, censor date was set to the date of death of non-CVD causes, or the last hospitalization date known for each patient.

### Statistical analyses

In our primary analyses focusing on the association between mCAs and death of CVD causes, CAD causes, from cancer, and death of any cause, Cox proportional hazards regression models were used, adjusting for age at baseline, sex, smoking status, treatment with chemotherapy and/or radiotherapy, the number of days between cancer diagnosis and date of baseline, and principal components for genetic ancestry (PC, 1-10). An mCA-by-cancer status interaction term was also assessed with respect to each of the primary endpoints in the overall population including individuals without any history of cancer (n=479 435). We further examined the associations of mCA types (autosomal mCAs, mosaic loss of the Y chromosome, mosaic loss of the X chromosome, and expanded mCAs) on the primary endpoints of interest. Sensitivity analyses considered the additional adjustment for hypertension and body mass index. Cox regression models were also used for exploratory analyses of incident CV phenotypes. Sub-group analyses were conducted according to cancer types, smoking status, treatment with chemotherapy, and age at baseline dichotomized at 65 years. All analyses were performed using the survival package in R (version 4.1.2, Bird Hippie). All analytical and summary reports were produced with gtsummary (version 1.6.1).^45^

## Supporting information

sFigure 1

## Data Availability

All data produced in the present study are available upon reasonable request to the authors

## Acknowledgements

We thank the UK Biobank for providing the data under Application Number 20168.

## Figure Legends

**sFigure 1**. Visual depiction of the proportion of patients with at least 1 mosaic chromosomal alteration in the overall cohort (A), of the proportion of patients with at least one mosaic chromosomal alteration stratified according to age groups (B), and of the proportion of patients with at least one mosaic chromosomal alteration stratified according to cancer types (C). mCA: mosaic chromosomal alteration.

