## Supplementary figures and images for "Somatic mosaic chromosomal alterations and death of cardiovascular disease causes among cancer survivors: an analysis of the UK Biobank"

### sFigure 1

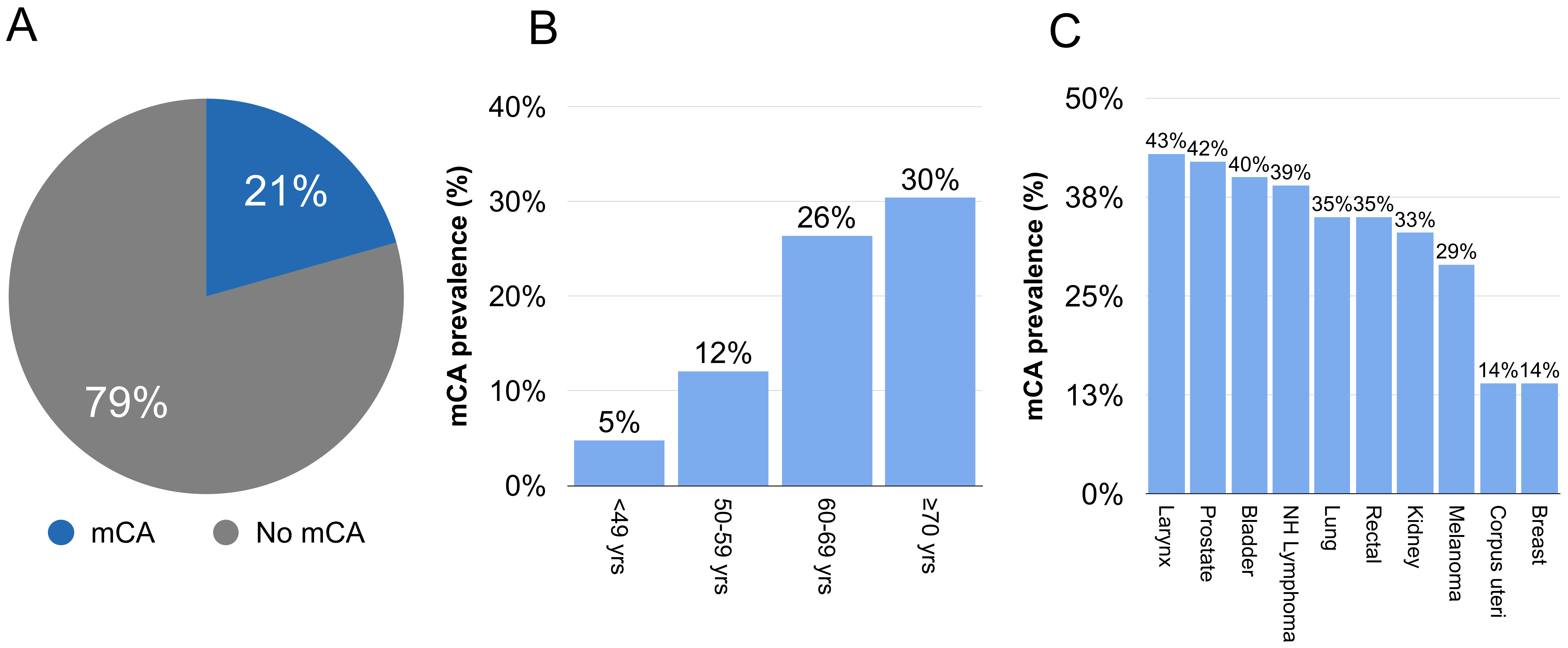
